# Early neurodevelopmental problems and risk for avoidant/restrictive food intake disorder (ARFID) in the general child population: a Japanese birth cohort study

**DOI:** 10.1101/2021.11.08.21265646

**Authors:** Lisa Dinkler, Kahoko Yasumitsu-Lovell, Masamitsu Eitoku, Mikiya Fujieda, Narufumi Suganuma, Yuhei Hatakenaka, Nouchine Hadjikhani, Rachel Bryant-Waugh, Maria Råstam, Christopher Gillberg

## Abstract

**Background:** An overrepresentation of neurodevelopmental disorders (NDDs) has been observed in individuals with avoidant/restrictive food intake disorder (ARFID). Previous studies on the association between ARFID and NDDs are limited to cross-sectional data from clinical samples of small size. This study aimed to extend previous research by using prospectively collected data in children from a general population sample. We examined the occurrence and predictive power of early neurodevelopmental problems in 4–7-year-old children with suspected ARFID.

**Methods:** Data were collected via parent-report in 3,728 children born between 2011 and 2014 in Kochi prefecture, a sub-sample of the Japan Environment and Children’s Study (JECS). Neurodevelopmental problems were assessed with several instruments at different time points between 0.5 and 3 years of age as part of the JECS. In an add-on study, ARFID was identified cross-sectionally (between 4 and 7 years of age) using a newly developed screening tool.

**Results:** Circa 3% of children at high risk for NDDs in preschool age screened positive for ARFID between age 4 and 7 years, reflecting a three times increased risk of suspected ARFID. A fifth (20.8%) of children with suspected ARFID had likely NDDs, compared to 8.6% of children without suspected ARFID. Developmental delay trajectories of children with and without suspected ARFID started to divert after the age of 6 months. Only 2.2% of children with early feeding problems later screened positive for ARFID. The inclusion of neurodevelopmental problems improved the prediction of later ARFID.

**Conclusions:** The results mirror the previously observed overrepresentation of NDDs in ARFID populations, although to a weaker extent. In non-clinical populations, early feeding problems are common and rarely develop into ARFID, however, our findings imply that they should be monitored closely in children with high neurodevelopmental risk in order to prevent ARFID.

## Introduction

Avoidant/restrictive food intake disorder (ARFID) is characterized by a persistent restriction of food intake in amount and/or variety that results in weight loss or failure to gain weight, insufficient growth, nutritional deficiency, dependence on enteral feeding or oral nutritional supplementation, and/or marked interference with psychosocial functioning (American Psychiatric Association, 2013). Contrary to other eating disorders such as anorexia nervosa and bulimia nervosa, ARFID is not motivated by body image concerns or drive for thinness. Instead, food avoidance/restriction in ARFID is often based on one or more of three “drivers”: (1) concern about aversive consequence of eating (e.g., choking, vomiting), (2) sensory-based avoidance (e.g., based on the smell, taste, appearance, or consistency/texture of foods, and (3) lack of interest in food or eating (American Psychiatric Association, 2013).

A higher than predicted occurrence of neurodevelopmental disorders (NDDs) has been observed in patients with ARFID compared to general population estimates, with some studies identifying higher rates of co-occurrence with ARFID than with anorexia nervosa (Nicely et al., 2014, Norris et al., 2021, Lieberman et al., 2019). In children and adolescents with ARFID seen in feeding/eating disorders services, the prevalence of specific NDDs has been estimated at 3-23% for autism spectrum disorder (ASD) (Kambanis et al., 2020, Reilly et al., 2019, Norris et al., 2021, Lieberman et al., 2019), 3-39% for attention-deficit/hyperactivity disorder (ADHD) (Reilly et al., 2019, Nicely et al., 2014, Norris et al., 2021, Duncombe Lowe et al., 2019, Lieberman et al., 2019), 10-31% for learning difficulties/disorders (Nicely et al., 2014, Lieberman et al., 2019, Norris et al., 2021), and 26-38% for intellectual disability or general developmental delay (Nicely et al., 2014, Sharp et al., 2020).

To the best of our knowledge, research on ARFID and NDDs has so far almost exclusively been of cross-sectional nature and limited to specific clinical samples from the US and Canada with small sample size. The only larger study estimated that in a cohort of 5,157 individuals with ASD—largely identified through clinical sites—21% were at high risk for ARFID (Koomar et al., 2021). Clinical samples might be biased in that children with higher severity, potentially caused by multi-comorbidity, might be overrepresented. Further, the recognition of the ARFID diagnosis is relatively new and referral routes are most often not yet standardized. Children with ARFID can therefore be encountered in a range of different specialties (e.g., paediatrics, psychiatry, gastroenterology, dietetics, occupational therapy). For instance, while children with ARFID and comorbid medical conditions might be referred to paediatric clinics, those with ARFID and considerable fear/anxiety might be referred to child and adolescent mental health services. The estimated prevalence of NDD comorbidity might therefore heavily depend on the specific speciality a sample was drawn from. In contrast, samples screened from the general population are potentially more representative of the entire group of individuals affected by ARFID, including those who are not seeking treatment. To date, no studies examining the association of ARFID and NDDs on the general population level exist.

It has been well-established that early neurodevelopmental symptoms are highly predictive of later diagnosed NDDs and aid in the early detection of children with NDDs, enabling early interventions (Gillberg, 2010). Feeding difficulties are considered one of these symptoms, and in children with ASD, feeding difficulties often constitute one of the first problems parents are worried about and seek help for (Barnevik Olsson et al., 2013). Due to the suggested significant overlap between ARFID and NDDs, and the fact that children with NDDs are not always diagnosed or often diagnosed very late (Gould and Ashton-Smith, 2011, Huang et al., 2020), the question arises whether early neurodevelopmental problems can also aid in the early detection of ARFID. This has not been examined using longitudinal data.

### Aim of this study

The present study extends previous research on ARFID and NDDs by using prospectively collected data in children from a general population sample. We aimed to examine the occurrence and predictive power of early neurodevelopmental problems in 4–7-year-old children with suspected ARFID. We investigated the following specific questions: (1) Are children with early neurodevelopmental problems at increased risk for ARFID and if so, how many of these children will have ARFID? (2) Which specific types of neurodevelopmental problems are predictive of ARFID? (3) When do children with ARFID on average start to divert in their neurodevelopmental development?

## Methods

### Study population

We conducted a cross-sectional parental survey in a sub-sample of the Japan Environment and Children’s Study (JECS), using a screening tool for ARFID recently developed by our group (Dinkler et al., 2021). JECS is an ongoing nationwide birth cohort study, investigating environmental factors affecting children’s health and development (Kawamoto et al., 2014, Michikawa et al., 2018). Our study was conducted in collaboration with the Kochi Regional Centre of JECS at Kochi Medical School. In December 2018, the survey was sent out to the parents of 6,633 JECS participants in Kochi prefecture, born between July 2011 and December 2014. The response rate was 56.5% (n=3,746). After excluding 18 children due to missing/unclear responses relating to ARFID criteria, the final sample consisted of 3,728 children. Attrition analyses showed that on average the children in this sample might be slightly healthier than the Japanese child population (see supplement in (Dinkler et al., 2021)). The study was approved by the Ethics Committee at Kochi Medical School (ERB-102925 and ERB-104083). Informed consent was obtained from all participants.

### Measurements

#### ARFID screening

ARFID was assessed using a newly developed, parent-reported screener for children aged 2 years and up. The development and contents of the ARFID screener are described in Dinkler et al. (2021). In short, the screener contains 10 items that closely map onto the DSM-5 diagnostic criteria for ARFID. The items and the screening algorithm are shown in Table S1. In addition, the three drivers of food avoidance/restriction in ARFID were measured with one item each (Table S1). For details regarding response options and required responses to meet the respective criterion please refer to Dinkler et al. (2021).

Children were identified as screening positive for ARFID when they met DSM-5 ARFID criterion A, plus at least one of criteria A1, A2, A3, and A4, as well as criteria C and D. Criterion B (the eating disturbance is not due to lack of available food or a culturally sanctioned practice) was not assessed as we considered our cohort (a) affluent enough for food shortage to be relatively unlikely, and (b) culturally homogenous enough with no particular food restriction practice. For reasons of readability, children screening positive for ARFID are referred to as *children with suspected ARFID*, and children screening negative for ARFID are referred to as *children without suspected ARFID* in the following.

#### Prospective assessment of neurodevelopmental problems (before ARFID screening)

Data on neurodevelopmental problems came from different sources (Figure 1). As part of the JECS main study, the Japanese version of the **Ages and Stages Questionnaire-3** (Squires and Bricker, 2009, J-ASQ-3; Mezawa et al., 2019) was collected every six months after birth. Data were available until the age of 3 years for the current study. The ASQ-3 assesses parent-reported developmental delay in five skill domains: communication (language skills), gross motor (e.g., sitting, crawling, walking, running), fine motor (hand and finger movement/coordination), problem-solving (e.g., playing with toys) and personal-social (self-help skills and interaction). Each domain consists of six questions on whether a certain activity can be done by the child, rated with “yes” (10 points), “sometimes” (5 points), or “not yet” (0 points). Two cut-off values exist for the resulting domain scores at each age to identify children potentially at risk for developmental delay: a “monitoring cut-off” at 1 standard deviation (SD) below the mean and a “referral cut-off” at 2 SD below the mean (Mezawa et al., 2019).

**Figure 1.**
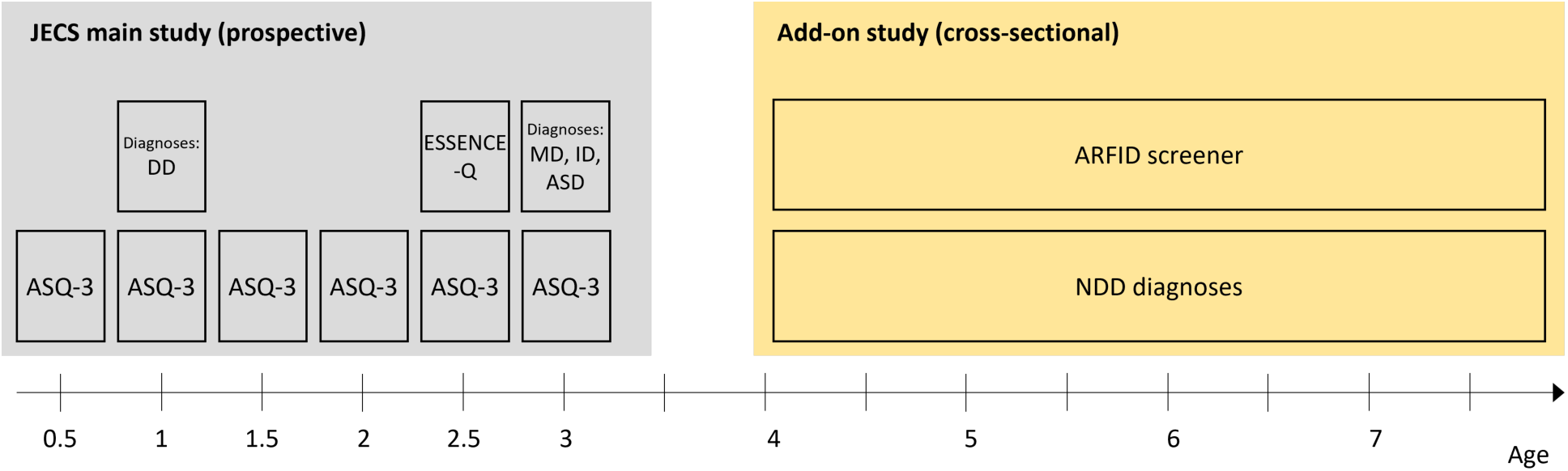
ASD: autism spectrum disorder, ASQ-3: Ages and Stages Questionnaire, DD: developmental delay, ESSENCE-Q: Early Symptomatic Syndromes Eliciting Neurodevelopmental Clinical Examinations-Questionnaire, ID: intellectual disability, MD: motor delay, NDD: neurodevelopmental disorder. All measures were parent-reported.

We also used data from the **Early Symptomatic Syndromes Eliciting Neurodevelopmental Clinical Examinations Questionnaire** (ESSENCE-Q; Hatakenaka et al., 2016), which was collected in the JECS main study at child age 2.5 years. The ESSENCE-Q screens for a broad range of early neurodevelopmental symptoms that might indicate the presence of NDDs and therefore suggest clinical examination. The ESSENCE-Q consists of 11 short questions which ask whether the parents or someone else has been concerned regarding a range of developmental areas for more than a few months. In the original ESSENCE-Q, the questions are rated with “yes”, “maybe/a little” or “no”. In the JECS, the response format was changed to “yes” or “no”.

In addition, parents were asked at child age 1 year whether their child had been diagnosed with developmental delay, and at child age 3 years whether their child had been diagnosed with motor delay, intellectual disability or ASD.

We derived a **neurodevelopmental risk score** by aggregating the above described measures as follows: scoring below the referral cut-off on an ASQ-3 domain—1 risk point (6 time points of measurement on 5 ASQ-3 domains: max. 30 points); ESSENCE-Q items except feeding concerns, rated with “yes”—1 risk point (max. 10 risk points); each indication of developmental delay, motor delay, intellectual disability or ASD at child age 1 or 3 years, respectively—1 risk point (max. 4 risk points). In total, the neurodevelopmental risk score had a theoretical range from 0 to 44. Feeding problems were not included into the neurodevelopmental risk score so we could examine to which degree cumulative neurodevelopmental risk predicted later ARFID over and above previous feeding problems.

To make use of the repeated measurements of the ASQ-3 between 0.5 and 3 years of age, we computed individual **ASQ-3 risk scores** for each time point of measurement as follows: scoring below the monitoring cut-off on an ASQ-3 domain—1 risk point; scoring below the referral cut-off on an ASQ-3 domain—2 risk points. Summed up over the five ASQ-3 domains, this yielded an individual ASQ-3 risk score with a theoretical range of 0-10 points per time point of measurement.

#### Cross-sectional assessment of NDD diagnoses (concurrent with ARFID screening)

In the survey that was sent out to parents at child age 4-7 years, parents were also asked to indicate whether their child had received a NDD diagnosis, including ASD, ADHD, developmental coordination disorder, intellectual disability, tic disorder/Tourette syndrome, specific learning disorder (e.g. dyslexia), oppositional defiant disorder, and conduct disorder.

#### Validity of the neurodevelopmental risk score

Apart from analysing the continuous neurodevelopmental risk score, we also created binary variables comparing those with highest neurodevelopmental risk (i.e., scoring above the 80^th^, 90^th^, 95^th^ and 99^th^ percentile) to those with lower neurodevelopmental risk. The prevalence of NDDs in the general child population is roughly 10% (Gillberg, 2010). Children scoring above the 90^th^ percentile on the neurodevelopmental risk score are, therefore, likely to approximately represent the population with NDDs. Similarly, those scoring above the 80^th^ percentile are likely at increased risk of NDDs, while those above 95^th^ and 99^th^ percentile will almost certainly have NDDs, probably even several ones. To investigate the validity of the neurodevelopmental risk score we calculated the odds of having any diagnosed NDD measured cross-sectionally at age 4-7 years for children scoring above the 80^th^, 90^th^, 95^th^ and 99^th^ percentiles of the neurodevelopmental risk score. Odds ratios were as follows: above 80^th^ percentile: OR=10.08 (95% CI 7.05-14.39); above 90^th^ percentile: OR=13.71 (95% CI 9.62-19.53); above 95^th^ percentile: OR=16.62 (95% CI 11.36-24.31); above 99^th^ percentile: OR=31.17 (95% CI 16.68-58.25). Although confidence intervals were large, these data show that the neurodevelopmental risk score used in this study is a good approximation of risk for NDDs.

#### Statistical analyses

Descriptive statistics were used to report sample characteristics and frequency of neurodevelopmental problems in children with versus without suspected ARFID. ARFID status (screen-positive vs. screen-negative for ARFID) was used as dependent variable throughout. We applied logistic regressions to calculate odds ratios for neurodevelopmental problems in children with versus without ARFID. To determine the most important domains within ESSENCE-Q and ASQ-3, we used stepwise multiple logistic regression with backward elimination (significance level for removal from the model: p>.20, significance level for addition to the model: p<.10). Stata 16.1 was used for data analysis (StataCorp, 2019) and R 4.0.0 for data visualization (R Core Team, 2020).

## Results

Demographic characteristics of the sample (n=3,728) are presented in Table 1. Almost all questionnaires were completed by mothers (98.1%). Forty-nine children (1.3%) were identified with ARFID (22 boys, 27 girls; see Dinkler et al. 2021). The frequency of specific diagnostic criteria in children with ARFID can be found in Table S1. Lack of interest in food or eating (63.3%) and sensory-based avoidance (51.0%) were the most common drivers of food avoidance, while concern about aversive consequences of eating was less common (14.3%).

**Table 1.**
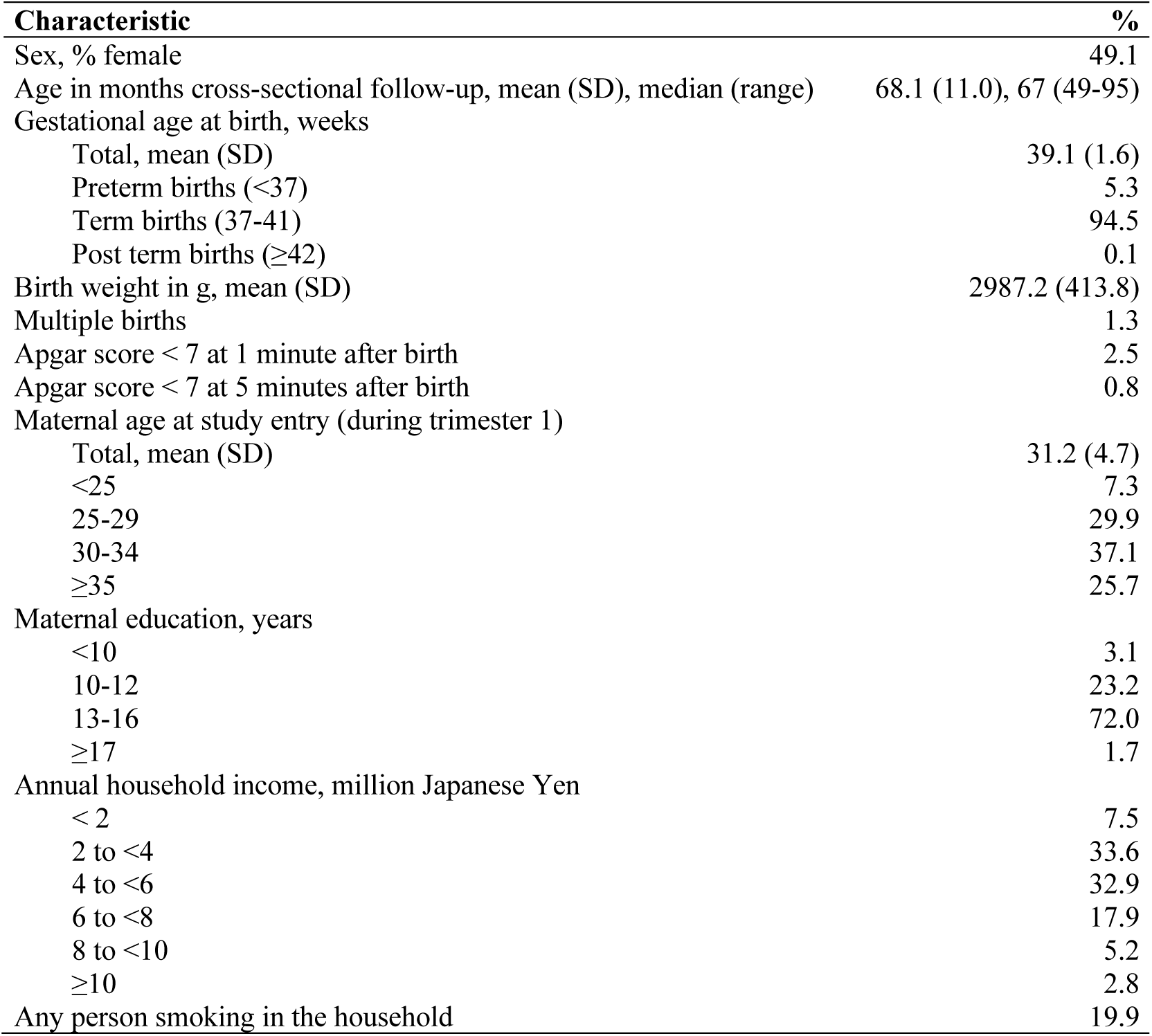
Demographic characteristics of the sample (n=3,728)

| Characteristic | % |
| --- | --- |
| Sex, % female | 49.1 |
| Age in months cross-sectional follow-up, mean (SD), median (range) | 68.1 (11.0), 67 (49-95) |
| Gestational age at birth, weeks |  |
| Total, mean (SD) | 39.1 (1.6) |
| Preterm births (<37) | 5.3 |
| Term births (37-41) | 94.5 |
| Post term births (≥42) | 0.1 |
| Birth weight in g, mean (SD) | 2987.2 (413.8) |
| Multiple births | 1.3 |
| Apgar score < 7 at 1 minute after birth | 2.5 |
| Apgar score < 7 at 5 minutes after birth | 0.8 |
| Maternal age at study entry (during trimester 1) |  |
| Total, mean (SD) | 31.2 (4.7) |
| <25 | 7.3 |
| 25-29 | 29.9 |
| 30-34 | 37.1 |
| ≥35 | 25.7 |
| Maternal education, years |  |
| <10 | 3.1 |
| 10-12 | 23.2 |
| 13-16 | 72.0 |
| ≥17 | 1.7 |
| Annual household income, million Japanese Yen |  |
| < 2 | 7.5 |
| 2 to <4 | 33.6 |
| 4 to <6 | 32.9 |
| 6 to <8 | 17.9 |
| 8 to <10 | 5.2 |
| ≥10 | 2.8 |
| Any person smoking in the household | 19.9 |

### Neurodevelopmental problems at age 0.5-3 years (prospectively)

#### Neurodevelopmental risk score and later ARFID

Analyses showed that the higher the neurodevelopmental risk score (including repeated measures between 0.5 and 3 years of age) the higher the risk for ARFID measured cross-sectionally when children were between 4 and 7 years of age. The odds for ARFID increased by 11% for each unit increase on the neurodevelopmental risk score (theoretical range: 0-44; Table 2). Children in the highest risk percentiles had roughly three times higher odds of having ARFID, except for those above the 99^th^ percentile, where the odds for having ARFID were 8.43, however with a broad confidence interval. The absolute risks of later ARFID for children above the 80^th^ (90^th^, 95^th^, 99^th^) percentile on the neurodevelopmental risk score were 2.5% (3.1%, 3.7%, 9.3%) compared to an ARFID prevalence of 1.3% in the total sample. A fifth (20.8%) of children with ARFID had a neurodevelopmental risk score above the 90^th^ percentile, indicating the presence of one or more NDDs, compared to 8.6% of children without ARFID (OR=2.80, 95% CI 1.38-5.67; Table 2).

**Table 2.**
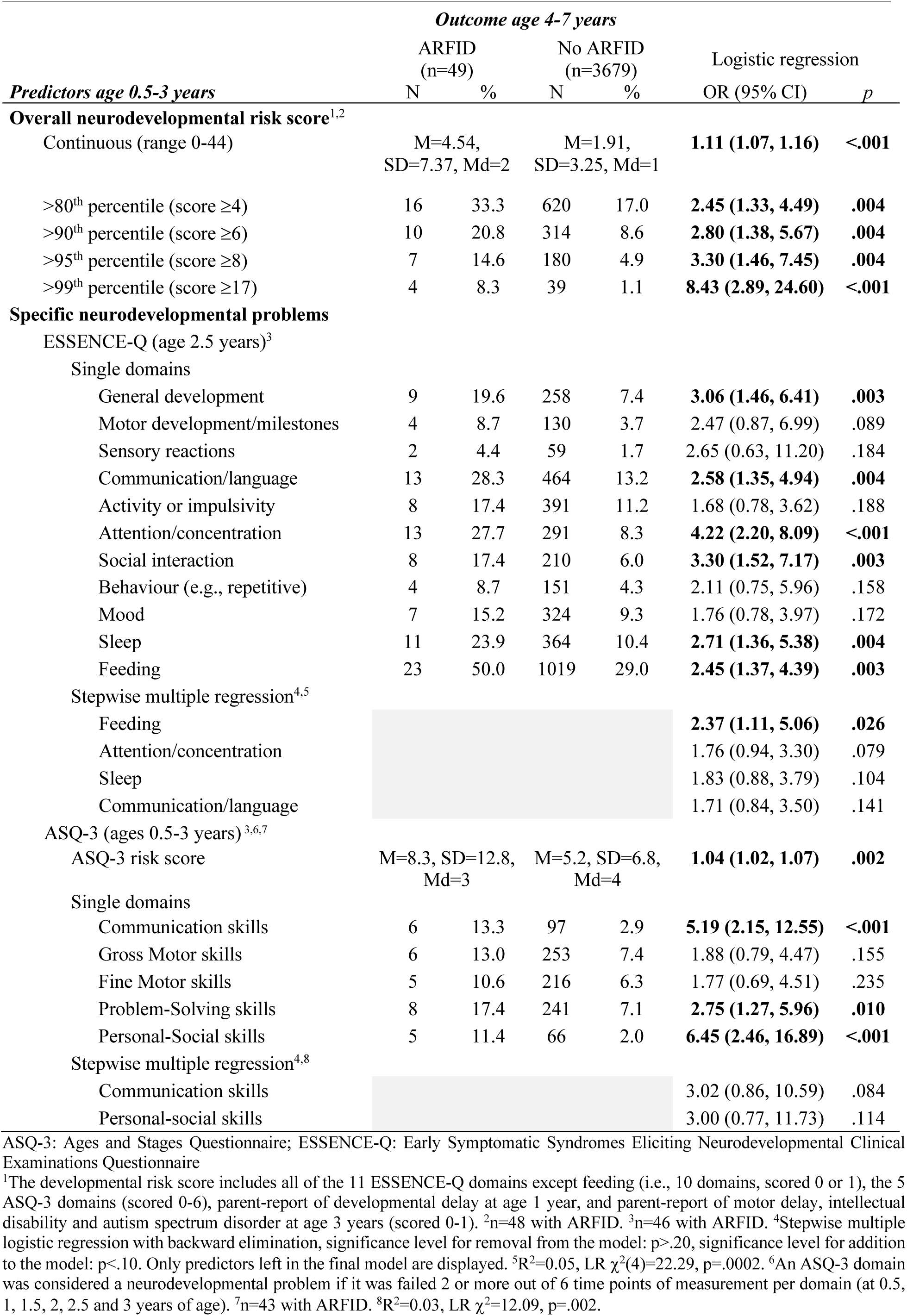
Longitudinal/prediction: Early neurodevelopmental problems/concerns/risk at age 0-3 years in children screening positive versus negative for ARFID at age 4-7 years

We ran logistic regressions to examine whether children with ARFID and *high* neurodevelopmental risk (>90^th^ percentile) differed from children with ARFID and *low* neurodevelopmental risk (<90^th^ percentile) in drivers of food avoidance. High neurodevelopmental risk was associated with a higher number of drivers (OR=1.42, 95% CI 1.23-1.65) owing to a higher presence of sensory-based food avoidance (OR=1.70, 95% CI 1.34-2.14) and lack-of-interest-based food avoidance (OR=1.53, 95% CI 1.15-2.03), while concern-based food avoidance was equally common (OR=1.18, 95% CI 0.78-1.77).

#### Specific predictors of later ARFID

Of the single ESSENCE-Q domains reported at age 2.5 years, the following were significantly associated with later ARFID in simple logistic regressions: general development, communication/language, attention/concentration, social interaction, sleep and feeding (OR range 2.45-4.22; Table 2). In the stepwise multiple logistic regression, four neurodevelopmental problems were retained in the final model: feeding, attention/concentration, sleep, and communication/language, with feeding being the only predictor of later ARFID that reached significance at the .05 level (Table 2). The multiple regression was relatively low-powered with only four events per variable (46 cases with ARFID divided by 11 candidate variables = 4.2). Early feeding problems were very common in this sample (29.3%), but only 2.2% of children with early with feeding problems (23 out of 1042) later screened positive for ARFID. Only 50% of children screening positive for ARFID had early feeding problems.

To investigate the single ASQ-3 domains, we considered whether a child scored below the referral cut-off at *two or more* (vs. at less than two) of the six time points of measurement between 0.5 and 3 years of age. The following domains were significantly associated with later ARFID in simple logistic regressions: communication skills, problem-solving skills, and personal-social skills (OR range 2.75-6.45; Table 2). In the stepwise multiple logistic regression, communication skills and personal-social skills were retained in the final model, but none of them reached significance at the .05 level. Again, power was relatively low with nine events per variable (46 cases with ARFID divided by 5 candidate variables = 9.2).

#### Prediction of later ARFID

We ran a logistic regression to test how much the inclusion of the whole range of neurodevelopmental problems assessed in this study (11 in ESSENCE-Q, 5 in ASQ-3) could improve prediction of later ARFID beyond early feeding problems. The model containing feeding problems-only explained 1.8% of the variance, while the full model explained 6.6% of the variance.

#### ASQ-3 risk score trajectories and later ARFID

Lastly, we investigated ASQ-3 risk score trajectories over the six time points of measurement between

0.5 and 3 years of age (theoretical range ASQ-3 risk score: 0-10 points per time point of measurement).

Both groups—children with and without suspected ARFID—started out at same level at 0.5 years. After the age of 0.5 years, the scores started diverting: while the ASQ-3 risk scores decreased and stayed at a low level in the group without suspected ARFID, scores stayed high in the ARFID group, indicating an early diversion of developmental trajectories (Figure 2).

**Figure 2.**
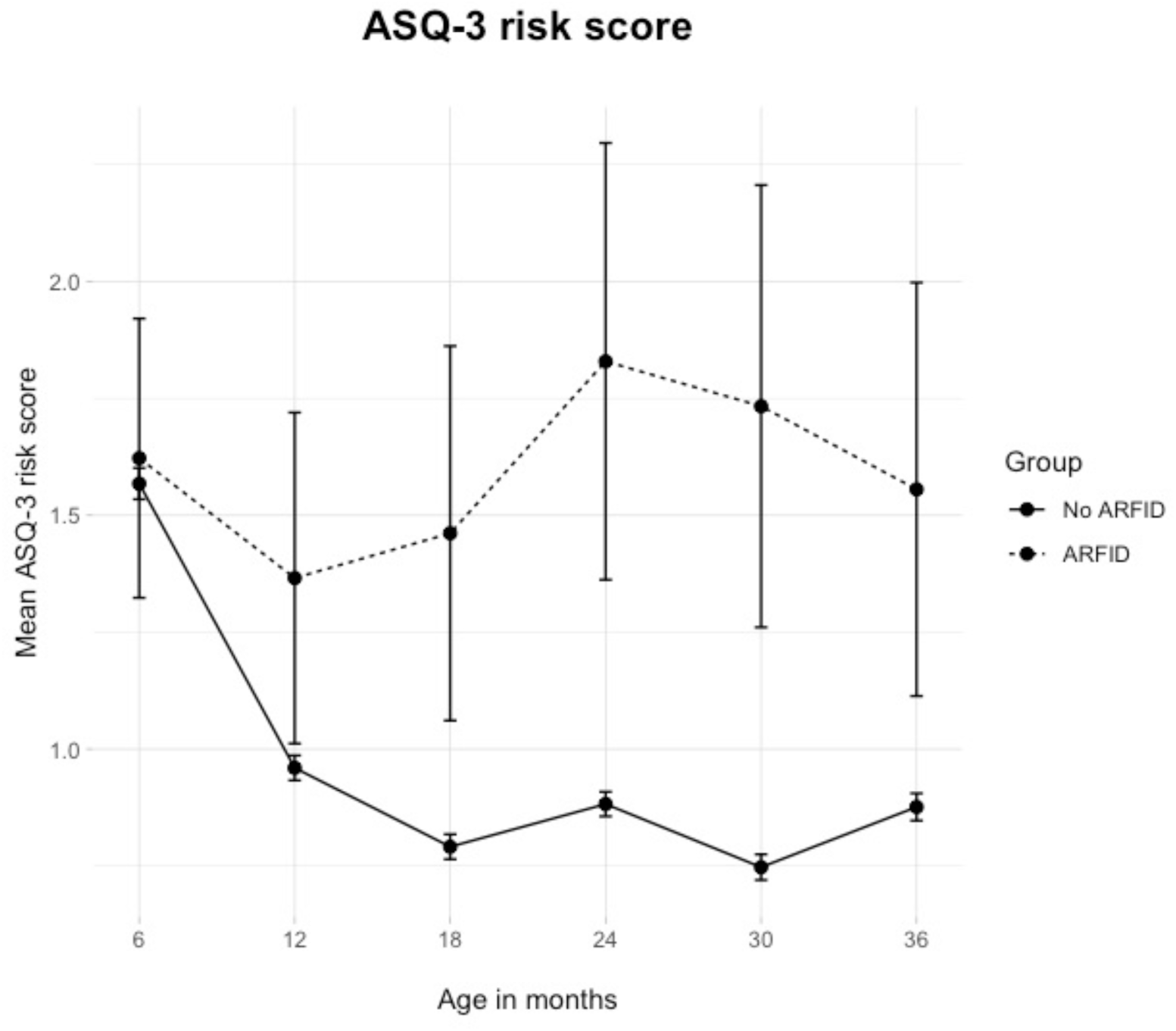
Mean Ages and Stages Questionnaire (ASQ-3) risk score across age by ARFID group. The x-axis represents age in months. The y-axis represents the mean ASQ-3 risk score in the ARFID group (dashed line) versus the no-ARFID group (solid line) with a theoretical range of 0-10 points per measurement point (below the monitoring cut-off on an ASQ-3 domain: 1 risk point; below the referral cut-off on an ASQ-3 domain: 2 risk points; aggregated over the five ASQ-3 domains). Error bars represent the standard error of the mean.

### NDD diagnoses at age 4-7 years (cross-sectionally)

Children in the ARFID group had a significantly increased presence of parent-reported diagnoses of ASD (OR=5.19, 95% CI 1.81-14.86) and intellectual disability (OR=12.00, 2.63-54.66; Table 3). 14.3% of children with ARFID had *any* of the NDD diagnoses included in the survey, compared to 3.6% of children without ARFID (OR=4.41, 95% CI 1.94-10.00). Power for these analyses was low due to the low frequency of ARFID and NDD diagnoses overall. Oppositional defiant disorder and conduct disorder were not present at all in this sample.

**Table 3.** Cross-sectional comorbidity with diagnosed neurodevelopmental disorders (NDD) in children screening positive versus negative for ARFID at age 4-7 years

|  | ARFID<br>(n=49) |  | No ARFID<br>(n=3679) |  | Logistic regression |  |
| --- | --- | --- | --- | --- | --- | --- |
|  | N | % | N | % | OR (95% CI) | p |
| Autism spectrum disorder | 4 | 8.2 | 62 | 1.7 | **5.19 (1.81, 14.86)** | .002 |
| ADHD | 1 | 2.0 | 49 | 1.3 | 1.54 (0.21, 11.41) | .671 |
| Developmental coordination disorder | 1 | 2.0 | 14 | 0.4 | 5.45 (0.70, 42.31) | .105 |
| Intellectual disability | 2 | 4.1 | 13 | 0.4 | **12.00 (2.63, 54.66)** | **.001** |
| Tic Disorders/Tourette Syndrome | 0 | 0.0 | 17 | 0.5 | ---- | ---- |
| Specific learning disorder (e.g., dyslexia) | 0 | 0.0 | 5 | 0.1 | ---- | ---- |
| Any NDD | 7 | 14.3 | 134 | 3.6 | **4.41 (1.94, 10.00)** | <.001 |

ADHD: Attention-Deficit/Hyperactivity Disorder

## Discussion

The present study sought to examine the occurrence and predictive power of early neurodevelopmental problems in 4–7-year-old children with suspected ARFID. We found that children at high risk for NDDs, measured between 0.5 and 3 years of age, were roughly three times more likely to screen positive for ARFID later on. This was confirmed by diagnoses of NDDs reported by parents at age 4–7 years, which were roughly four times more common in children with suspected ARFID. In absolute numbers, circa 3.1% of children scoring in the highest risk decile for NDDs developed ARFID, as opposed to 1.1% of children scoring below the highest risk decile for NDDs. Considered vice versa, circa 20.8% of children with suspected ARFID had likely NDDs, compared to 8.6% of children without suspected ARFID. ASQ-3 risk score trajectories over the six time points of measurement showed an early diversion of developmental trajectories (after the age of 6 months) in children with suspected ARFID.

Compared to previous studies which focused on the cross-sectional comorbidity of ARFID with NDDs, this study examined early neurodevelopmental symptoms in children with later suspected ARFID. Results confirmed the previously observed association between ARFID and NDDs, although the associations found in this sample from the general population were rather located at lower end of the ranges of previously reported estimates (see paragraph 2 in the introduction), which was in line with our expectations. Since previous studies from clinical samples reported estimates for *diagnosed* NDDs, our estimates are not directly comparable. We can, however, speculate that individuals with higher comorbidity might have a higher clinical severity leading to treatment-seeking and their inclusion into studies on clinical samples. For instance, in this sample, children with suspected ARFID and *high* neurodevelopmental risk on average had a higher number of drivers of food avoidance than children with suspected ARFID and *low* neurodevelopmental risk, potentially indicating a higher severity of ARFID. In addition, as a previous attrition analysis showed, our sample might have been slightly healthier than the average Japanese population, further weakening the detectable association between ARFID and neurodevelopmental problems. Alternatively, we may have underestimated the presence of neurodevelopmental problems, which was exclusively reported by parents, since the cohort was very young and neurodevelopmental problems sometimes do not become obvious before later childhood or adolescence (Mandy et al., 2018, Hosozawa et al., 2020).

Several specific early neurodevelopmental problems were associated with later ARFID in simple regression models, including symptoms of ASD (communication/language, social interaction) and ADHD (attention/concentration) as well as problems relating to general development, sleep and feeding. This is in line with previous research reporting an overrepresentation of ASD, ADHD and developmental delay in ARFID (Nicely et al., 2014, Lieberman et al., 2019, Reilly et al., 2019, Norris et al., 2021), as well as with the overrepresentation of ASD and intellectual disability diagnoses at follow-up in the present sample (while the present sample was largely too young for ADHD diagnoses, as evidenced by the low prevalence of 1.3%). However, in multiple regression models, none of the neurodevelopmental symptoms except feeding problems reached significance at the .05 level in predicting ARFID, which might be explained by the low power in these analyses due to the low number of children with ARFID in this sample. Surprisingly, early problems with sensory reactions were not significantly associated with later ARFID. Considering that 51% of the children with suspected ARFID showed sensory-based food avoidance, we would have expected to see increased problems with sensory reactions early on. The observed non-significant association could potentially be explained by the following. The ESSENCE-Q item for concerns around sensory reactions is very unspecific; it inquires hyper-as well as hyposensitivity to all kinds of sensory impressions (“e.g., touch, sound, light, smell, taste, heat, cold, pain”). Furthermore, sensory sensitivity to food characteristics might only in combination with low tolerance of variation/surprise and high level of risk avoidance lead to extremely cautious eating behaviour (i.e., food avoidance).

In line with previous research on eating behaviour in early childhood (Taylor et al., 2015), feeding problems at age 2.5 years were very common (29.3% of all children). However, only 2.2% of the children with early feeding problems were later suspected of having ARFID, which shows that early feeding problems as reported by parents in the ESSENCE-Q are unspecific and not a good predictor of later ARFID. Our results also showed that this prediction could be significantly improved by including the whole range of neurodevelopmental problems assessed in this study. Vice versa, only 50% of children screening positive for ARFID had early feeding problems, indicating that in half of the children, the onset of ARFID might have been after the age of 2.5 years.

### Strengths and limitations

Our study has several strengths. To our knowledge, this is the first study to examine prospectively collected data on a broad range of early neurodevelopmental problems in children screening positive for ARFID in a large general population sample. This enabled us to study the occurrence and predictive power of early neurodevelopmental problems in an ARFID group that is likely to be representative of 4-7-year-old Japanese children with ARFID, as it also includes those who do not seek treatment and hence are not part of clinical samples.

Several limitations must be considered. First, in the present study we screened for ARFID using a newly developed tool which still has to be validated against clinical ARFID diagnoses. In a previous study using this screening tool we found promising initial evidence of convergent validity with a range of measures assessing restrictive type eating as well as with weight and height (Dinkler et al., 2021). Second, although the total sample was relatively large, the number of children with ARFID was low, resulting in low power for the multiple regression analyses. Future research studying ARFID in the general population needs to employ even larger samples. Third, information on NDDs was collected through parent-reports only, although some of the questions asked about NDD *diagnoses*. Parents might not always be aware of neurodevelopmental symptoms or they might underreport diagnoses due to associated stigma. On the other hand, parents might be over-worried and therefore overreport symptoms, whereas overreporting of diagnoses seems unlikely. Optimally, neurodevelopmental symptoms and diagnoses should be clinically ascertained, which is, however, not always feasible, especially in epidemiological research including large samples like this one. Lastly, previous attrition analyses showed that our sample was slightly healthier than the average Japanese child population (Dinkler et al., 2021). It is therefore possible that children with ARFID in general, and specifically those with a higher disease burden or comorbid NDDs, were less likely to be included in our sample, which might have resulted in an underestimation of the association between ARFID and early neurodevelopmental problems, while an overestimation is unlikely.

## Conclusion

The present study showed that circa 3% of the children at high risk for NDDs in preschool age screened positive for ARFID later on (between age 4 and 7 years), which made them three times more likely to have suspected ARFID than children at low neurodevelopmental risk. Specific neurodevelopmental symptoms that were present at a higher rate in children with suspected ARFID reflected the increased prevalence of ASD, ADHD, developmental delay, and intellectual disability observed in clinical samples with ARFID. Our results largely mirror the previously reported overrepresentation of NDDs in individuals with ARFID, although our associations were less strong than some of the associations previously reported in clinical samples, which might include more severe cases with higher comorbidity. Considering the early onset of ARFID in many individuals and the observed comorbidity with NDDs, future research should investigate whether ARFID itself can be considered part of the NDD spectrum, at least in those with early onset and high NDD comorbidity. Our results also imply that, while early feeding problems are common and rarely develop into ARFID, they should be monitored closely in children with high neurodevelopmental risk, which can easily be screened for. In doing so, we might be able to prevent the worsening of eating pathology until full criteria for ARFID are met.

## Data Availability

Data are unsuitable for public deposition due to ethical restrictions and legal framework of Japan. It is prohibited by the Act on the Protection of Personal Information (Act No. 57 of 30 May 2003, amendment on 9 September 2015) to publicly deposit the data containing personal information. Ethical Guidelines for Medical and Health Research Involving Human Subjects enforced by the Japan Ministry of Education, Culture, Sports, Science and Technology and the Ministry of Health, Labour and Welfare also restricts the open sharing of the epidemiologic data. All inquiries about access to data should be sent to. The person responsible for handling enquiries sent to this e-mail address is Dr Shoji F. Nakayama, JECS Programme Office, National Institute for Environmental Studies.

## Abbreviations

ADHD: Attention-Deficit/Hyperactivity Disorder
ASD: Autism Spectrum Disorder
ARFID: Avoidant/Restrictive Food Intake Disorder
ESSENCE-Q: Early Symptomatic Syndromes Eliciting Neurodevelopmental Clinical Examinations-Questionnaire
ASQ-3: Ages and Stages Questionnaire-3
JECS: Japan Environment and Children’s Study
NDD: Neurodevelopmental disorder

## Acknowledgements

We would like to express our gratitude to all of the JECS study participants in the Kochi cohort and to the staff members at Kochi Regional Centre, who sent out and collected the questionnaires. This work was supported by the Swedish Research Council (M.R., 2018-02544; C.G., 538-2013-8864), Torsten Söderbergs Foundation (C.G., M151/14), AnnMari and Per Ahlqvist Foundation (C.G., 2018), Japan Society for the Promotion of Science (M.F., 18KK0263), Scandinavia-Japan Sasakawa Foundation (L.D., 2016), Samariten Foundation (L.D., 2017-0283), Wilhelm and Martina Lundgrens Foundation (L.D., 2017-1738), Petter Silfverskiölds Memorial Foundation (L.D., 2017-093 & 2018-142), Professor Bror Gadelius Memorial Foundation (L.D., 2019 & 2020), and Solstickan Foundation (L.D., 2020). The Japan Environment and Children’s Study was funded by the Ministry of the Environment, Japan. The funding sources had no role in the design and conduct of the study; collection, management, analysis, and interpretation of the data; preparation, review, or approval of the manuscript; and decision to submit the manuscript for publication. The findings and conclusions of this article are solely the responsibility of the authors.

## Conflict of Interest

The authors have no conflict to declare.

## Notes

### Competing Interest Statement

The authors have declared no competing interest.

### Funding Statement

This work was supported by the Swedish Research Council (M.R., 2018-02544; C.G., 538-2013-8864), Torsten Soederbergs Foundation (C.G., M151/14), AnnMari and Per Ahlqvist Foundation (C.G., 2018), Japan Society for the Promotion of Science (M.F., 18KK0263), Scandinavia-Japan Sasakawa Foundation (L.D., 2016), Samariten Foundation (L.D., 2017-0283), Wilhelm and Martina Lundgrens Foundation (L.D., 2017-1738), Petter Silfverskioelds Memorial Foundation (L.D., 2017-093 & 2018-142), Professor Bror Gadelius Memorial Foundation (L.D., 2019 & 2020), and Solstickan Foundation (L.D., 2020). The Japan Environment and Children's Study was funded by the Ministry of the Environment, Japan. The funding sources had no role in the design and conduct of the study; collection, management, analysis, and interpretation of the data; preparation, review, or approval of the manuscript; and decision to submit the manuscript for publication. The findings and conclusions of this article are solely the responsibility of the authors.

### Author Declarations

The study was approved by the Ethics Committee at Kochi Medical School (ERB-102925 and ERB-104083).

