## Supplemental Table 1 for "Early neurodevelopmental problems and risk for avoidant/restrictive food intake disorder (ARFID) in the general child population: a Japanese birth cohort study"

**Table S1** *Items used to screen the DSM-5 diagnostic criteria for avoidant/restrictive food intake disorder (ARFID), including the prevalence of criteria A1-A4 and drivers of food avoidance in the ARFID group*

| Criterion | Item | Prevalence in ARFID group in % |
|---|---|---|
| A - Avoidance or restriction of food intake | Do you think your child has an eating or feeding disturbance characterized by avoidance or restriction of food intake? (avoidance and restriction can relate to the range of foods eaten as well as the overall amount eaten) | 100.0 |
| A1 - Significant weight loss (or failure to grow/gain weight) | Over the past 3 months has there been concern that your child has not gained weight or grown as he/she should? | 49.0 |
| A2 - Significant nutritional deficiency | Has your child been identified by a health professional as having any nutritional deficiency? | 4.1 |
| A3 - Dependence on enteral feeding or oral nutritional supplements | a) Has your child been prescribed dietary supplements (e.g., vitamins, minerals) to address nutritional deficiencies? | 6.1 |
| | b) Did your child ever need nutritional supplement drinks (or other high-energy drinks) to be able to maintain/gain weight? | |
| A4 - Marked interference with psychosocial functioning | a) Do you believe that your child's *current* eating pattern causes any distress for your child? | 63.3 |
| | b) Does your child's *current* eating pattern interfere with his/her social functioning (e.g., attending preschool, affecting meals in preschool, making friends, play, activities)? | |
| C - Eating disturbance not attributable to weight/shape concerns | My child says that he/she feels fat, even if other people do not agree with him/her. (*item from Eating Disorder in Youth-Questionnaire, EDY-Q*) | ---- |
| D – Eating disturbance not attributable to concurrent medical condition | If your child has any problems with weight, growth or nutrition, is this primarily due to a current medical problem? Medical problem: (specify) | ---- |
| Driver - Lack of interest in food or eating | My child enjoys eating. *(item from Behavioral Pediatric Feeding Assessment Scale, BPFAS; reverse item)* | 63.3 |
| Driver – Sensory-based avoidance | My child dislikes to eat food with a specific smell, taste, appearance, temperature, or a certain consistency/texture (e.g., crispy or soft). | 51.0 |
| Driver – Concern about aversive consequences of eating | My child is afraid of eating because of worries about what might happen (e.g., choking, vomiting, stomach aches, diarrhoea, or allergic reactions etc.). | 14.3 |

**Screening algorithm:** A + (A1 or A2 or A3-a or A3-b or A4-a or A4-b) + C + D
